# A novel RNA nucleic acid amplification test more accurately distinguishes active *Clostridioides difficile* infection from colonization

**DOI:** 10.64898/2026.02.05.25342495

**Authors:** Kenneth D. Long, Daniel J. Silberger, John Hernandez, Thomas Detchemendy, Derek B. Moates, Tarek Abdalla, Lindsey E. Hastings, Aditi Jani, Janet Kim, Myles Prados, Melissa B. Miller, J. Martin Rodriguez, Sixto M. Leal

## Abstract

*Clostridioides difficile (C. diff)* is a leading cause of hospital-acquired infections with severity ranging from mild diarrhea to fulminant colitis and death. Current antigen tests lack adequate sensitivity and DNA-based nucleic acid amplification tests (DNA-NAATs) exhibit limited specificity for active infection, leading to either underdiagnosis or inappropriate treatment of colonized individuals. Unlike DNA, mRNA is expressed only by metabolically active bacteria and is rapidly hydrolyzed, providing natural advantages to distinguish active infection from colonization. In this study, we developed and evaluated a novel multiplexed reverse-transcriptase PCR assay (RNA-NAAT) targeting *C. diff*-specific sequences. No cross-reactivity was observed with other gastrointestinal pathogens, commensal enteric flora, or closely-related *Clostridioides* or *Clostridium* species. Toxin gene RNA expression was detected only in samples spiked with metabolically active *C. diff* at a limit of detection 30-to-50-fold less than current diagnostic methods. Sequential clinical samples (n=260) were collected in proprietary RNA-preservative solution from patients receiving standard of care *C. diff* diagnostic testing. All samples were evaluated with RNA-NAAT, Alere GDH/toxin EIA, Solana DNA-NAAT, and both forward- and reverse-2-Step Algorithms (2-SA). Samples yielding valid results on all platforms (n=239) with discordant test results (n=14) were adjudicated via toxigenic culture and blinded chart review by infectious disease physicians. RNA-NAAT outperformed all comparator test strategies, simultaneously exhibiting higher sensitivity and specificity, including a higher specificity for active infection than the specific toxin EIA (99.5% vs 98.2%) and a higher sensitivity for organism identification than the sensitive DNA-NAAT (100% vs 88.2%), with significantly reduced false positive test results (1 vs 7).

**One Sentence Summary:** A novel RNA diagnostic distinguishes clinically-relevant *C. difficile* infection from toxigenic carrier states, improving sensitivity and specificity.

## INTRODUCTION

*Clostridioides difficile* (*C. diff*) is a leading cause of hospital-acquired infections with severity ranging from mild diarrhea to fulminant colitis and death (*1-3*). *C. diff* diagnostics face multiple challenges. Enzyme immunoassays (EIAs) detecting Toxin A (TcdA) and Toxin B (TcdB) exhibit limited sensitivity, while EIAs targeting glutamate dehydrogenase (GDH) have low specificity (*4, 5*). DNA-based nucleic acid amplification tests (NAATs) targeting toxin-encoding genes exhibit increased analytical sensitivity but provide suboptimal specificity given their inability to distinguish toxigenic spores (colonization) from vegetative bacilli (active infection) (*6, 7*). With estimated rates of 10-30% colonization in hospitalized patients (*7, 8*), the limited specificity of DNA NAATs results in many false positive test results (*9-12*). Colonized individuals are then inappropriately treated with antibiotics resulting in further propagation of microbial dysbiosis, selective pressure for the development of antimicrobial resistance, and failure to identify the underlying cause of diarrheal illness (*13, 14*).

To address these diagnostic shortcomings, some institutions utilize two-step algorithms (2-SAs) (*15, 16*). In the forward 2-SA, GDH and Toxin EIAs are performed. Concordant positive and negative EIA results are reported without further testing. However, GDH^+^/Toxin EIA^-^ samples are reflex adjudicated via DNA NAAT. In the reverse 2-SA, the DNA NAAT is performed first with all negative results reported as final. Positive results undergo reflex adjudication with Toxin EIA. Overall, the forward 2-SA exhibits relatively low specificity and positive predictive value (resulting in false positives) (*13, 17*), while the reverse 2-SA exhibits reduced sensitivity (resulting in false negatives) (*18, 19*).

In this study, we developed and evaluated a novel RNA-based NAAT aimed at distinguishing metabolically inactive spores from vegetative cells, thereby better differentiating between active *C. diff* infection (CDI) and colonization. In addition to standard toxin gene targets used by DNA-NAAT (tcdA and tcdB), we looked at a highly-abundant 23S ribosomal RNA target (*10*), as well as binary toxin (cdtB) which has been associated with more severe clinical disease in prior literature (*20-22*). **Figure 1** highlights the diagnostic targets of this RNA NAAT and the rationale behind its ability to distinguish active CDI from colonization.

**Fig. 1.**
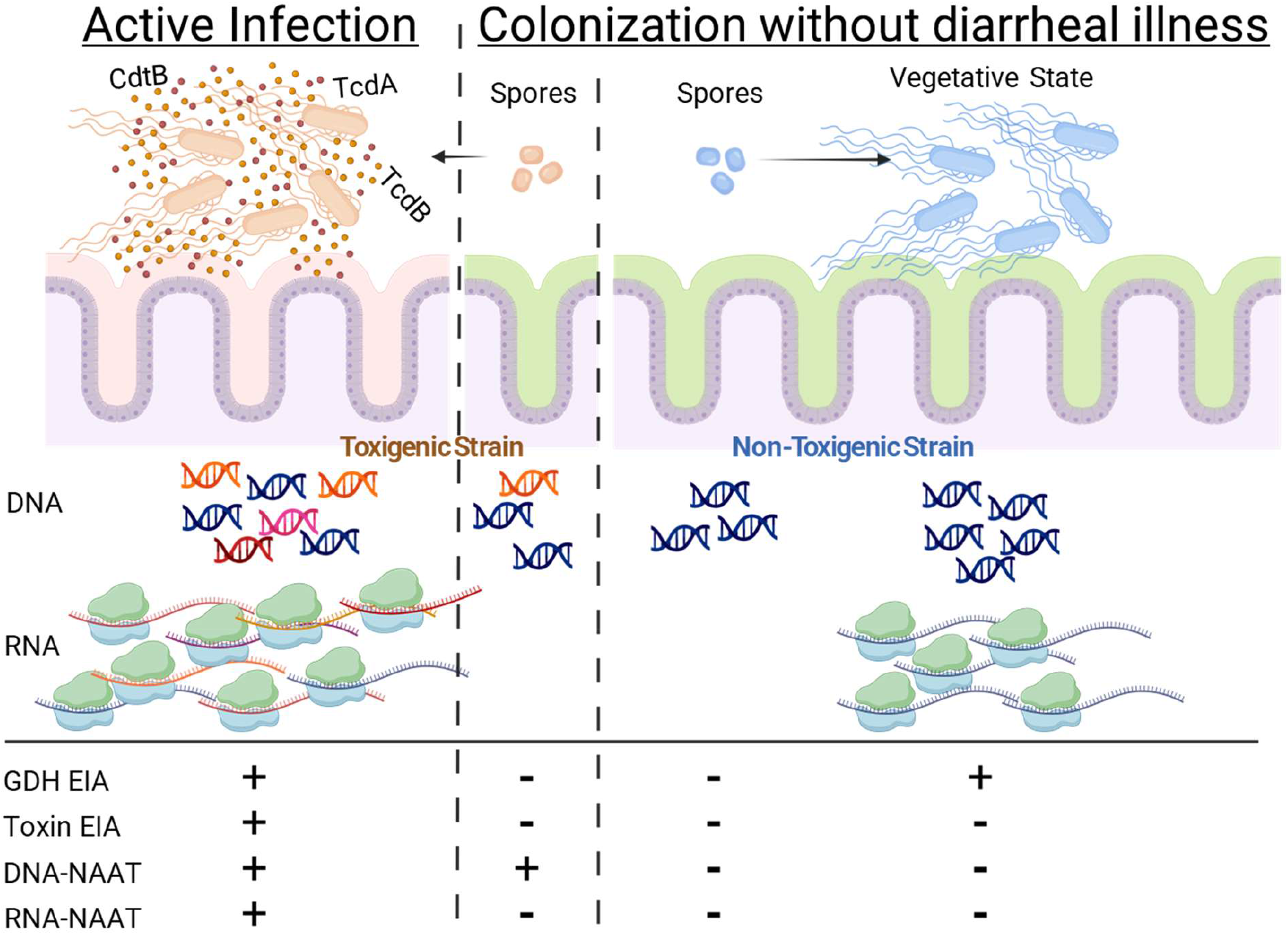
Diagnostic targets during active infection and colonization. Patients actively infected with toxigenic *C. diff* express GDH, TcdA, and TcdB and are expected to test positive for GDH and toxin EIAs, DNA-NAAT, and RNA-NAAT, with or without the presence of cdtB. In contrast, patients colonized with inactive *C. diff* spores that harbor toxin genes who are sick with diarrheal illness due to another cause would be expected to test positive with a GDH EIA and DNA NAAT but negative by toxin EIA and RNA NAAT. Patients with spores or active growth by nontoxigenic strains of *C. diff* express GDH and are expected to test positive by GDH EIA and negative for the 3 other tests. Based on these patterns, we hypothesized that the development of an RNA NAAT would enable the accurate diagnosis of active *C. diff* infection with improved specificity relative to DNA NAATs and increased sensitivity compared to toxin EIA.

## RESULTS

### RNA NAAT detects toxin RNA transcripts in metabolically active toxigenic *C. difficile*

To determine whether the primers in the RNA NAAT amplified a single target, we performed the assay on RNA extracts from a pure culture of metabolically active, toxigenic *C. diff* using SYBR Green dye (rather than target-specific probes). Each target—23S rRNA, tcdA, tcdB, and cdtB— produced a single peak, indicating specific amplification (**Figure 2A**). To determine the limit of detection (LOD) for each primer/probe set, defined as the minimum concentration with amplification above the threshold, we conducted the RNA NAAT assay on serial dilutions of toxigenic *C. diff* spiked into a PCR-negative stool matrix (**Figure 2B**). These data indicate an LOD of 150 CFU/mL (equivalent to 240 CFU/g of stool) for 23S rRNA (**Figure 2C**), 150,000 CFU/mL for *tcdA/B* (**Figure 2D**), and 150,000 CFU/mL for *cdtB* (**Figure 2E**).

**Fig. 2.**
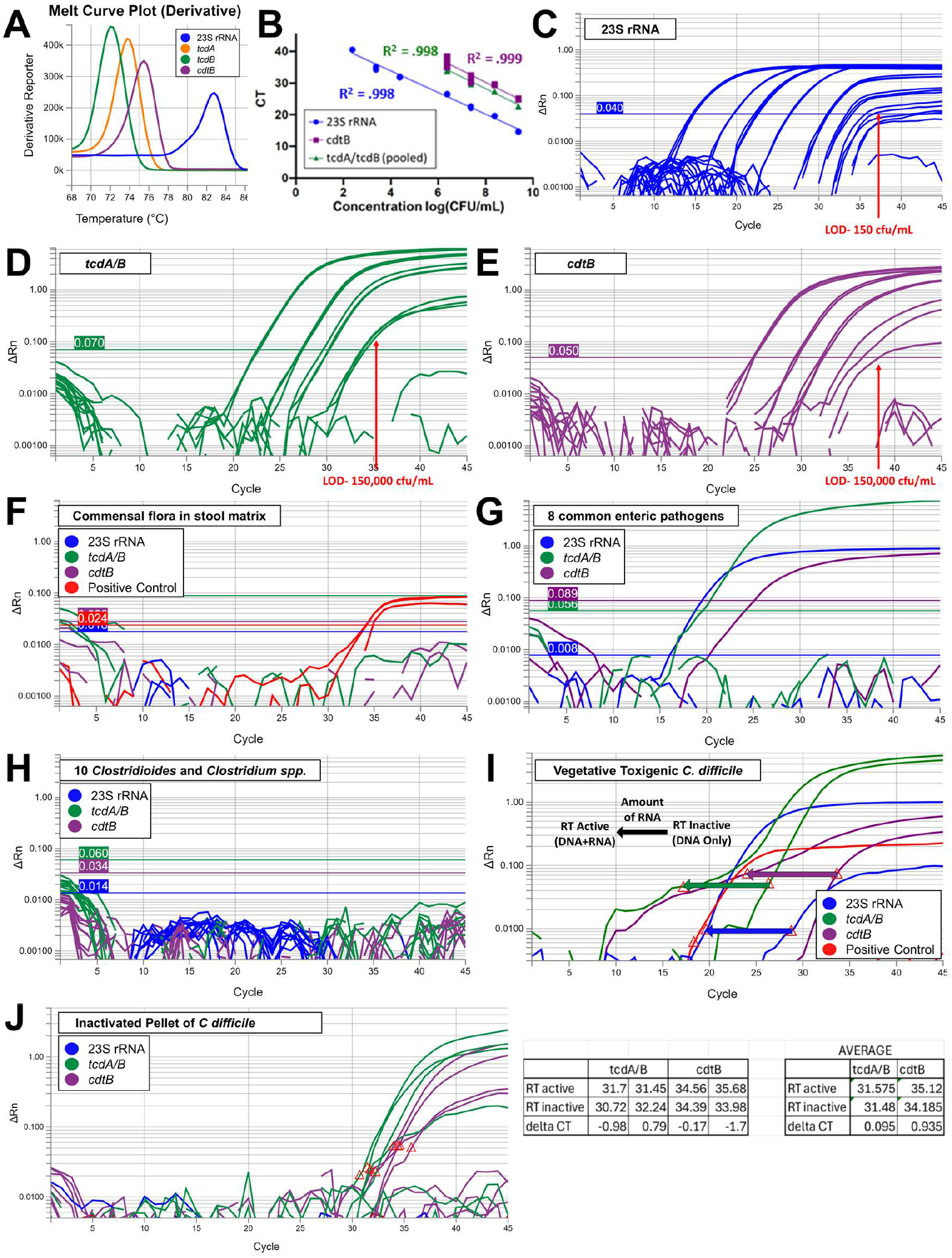
RNA NAAT analytical performance. **(A)** SYBR green melt curve analysis of species-specific primers detecting the *C. diff* 23s rRNA gene, as well as genes encoding TcdA, TcdB, and CdtB toxins. (**B)** Linearity of primer/probe sets amplifying 23S rRNA (VIC), *tcdA/tcdB* (FAM), and *cdtB* (TAMRA) from RNA extracts of toxigenic *C. diff* strain ATCC 9689. (**C)** Amplification curves indicating the limit of detection of the 23S rRNA gene, **(D)** *tcdA/B*, and (**E)** *cdtB* genes on RNA isolated from strain ATCC 9689. (**F)** RNA NAAT analysis showing no cross reactivity on RNA extracts from commensal flora in *C. diff* negative stool. (**G)** No cross-reactivity was observed using RNA-NAAT on 8 common enteric pathogens (*Vibrio parahaemolyticus, Campylobacter jejuni, Esherichia coli*, norovirus, *Shigella sonnei, Salmonella typhimurium, Yersinia enterocolitica, and* rotavirus, or (**H)** a panel of other *Clostridioides* and *Clostridium* species including *C. argentinense, C. baratti, C. bifermentans, C. clostridioforme, C. perfringens, C. ramosum, C. sordellii, C. sporogenes, C. tertium*, and *C. subterminale*. Amplification was observed with the positive control. **(I)** Amplification plot highlighting significant ΔCt shifts for 23S rRNA, *tcdA/B*, and *cdtB* upon heat inactivation of reverse transcriptase in the RT-PCR reaction using total nucleic acid from metabolically active toxigenic *C. diff*. (**J)** Amplification plot showing negligible ΔCt shifts in these targets using total nucleic acid from non-vegetative toxigenic *C. diff* (carrier state).

### RNA NAAT does not cross react with stool flora or enteric pathogens

To confirm that the primers/probes in the RNA NAAT are specific to *C. diff*, we evaluated nucleic acid extracts from commensal flora in stool matrix and observed no amplification/cross reactivity (**Figure 2F**). Additionally, cross reactivity was not observed with nucleic acid extracts from 8 common enteric pathogens (*Vibrio parahaemolyticus, Campylobacter jejuni, Escherichia coli, Shigella sonnei, Salmonella typhimurium, Yersinia enterocolitica*, norovirus, and rotavirus (**Figure 2G**), or closely related *Clostridium* and *Clostridioides* species, including *C. argentinense, C. baratti, C. bifermentans, C. clostridioforme, C. perfringens, C. ramosum, C. sordellii, C. sporogenes, C. tertium*, and *C. subterminale* (**Figure 2H**).

### Heat inactivation enables detection of toxin RNA expression from metabolically active toxigenic *C. difficile*

To determine if paired analysis of RT-PCR reactions, with or without heat inactivation of the reverse transcriptase enzyme, would result in an upward shift in the Ct value of the sample receiving heat inactivation (indicative of target RNA), we performed RNA NAAT on RNA extracts from a pure culture of metabolically active *C. diff* (**Figure 2I**) . Utilizing RNA extracts from toxigenic *C. diff*, each target—23S rRNA (blue), toxin A/toxin B (green), and binary toxin (purple) yielded an upward shift in the Ct value upon heat inactivation of 9.1, 9.1, 9.6, respectively (**Figure 2I**). This result is interpreted as **1**. Positive for *C. diff* organism based on 23S rRNA target positivity, and **2**. Positive for toxin RNA transcripts. This combination indicates that *C. diff* is present in the sample, and it is also actively transcribing the toxin genes that mediate tissue pathology. In contrast, a similar approach utilizing an already-inactivated (irradiation and heat) DNA extract from toxigenic *C. diff* showed no shift in Ct values for toxin genes upon reverse transcriptase heat inactivation (**Figure 2J**) (*23*).

### Receiver Operator Characteristic (ROC) curves identify optimal RNA NAAT performance at a delta Ct value threshold > 1.5

To identify an optimal RNA NAAT ΔCt value threshold, we evaluated 239 patient samples with all 6 test strategies and adjudicated discordant results via blinded chart review (**Figure 3). Figure 4** outlines the standard of care forward 2-SA utilized at our institution. Importantly, in this initial evaluation, RNA NAAT toxin A/B Ct shifts were recorded but not used to interpret results. Instead, toxin A/B target detection was the sole metric used to interpret RNA NAAT results. Utilizing this approach, 225/239 (94.1%) samples were concordant across all test platforms and 14/239 (5.9%) were discordant. Blinded chart review was used for definitive adjudication with 10/14 (71%) deemed negative and 4/14 (29%) deemed positive for active infection with toxigenic *C. diff*. Toxigenic culture was also performed, but it does not distinguish between metabolically inactive spores and vegetative bacilli and was therefore not used for adjudication. With this caveat, culture results showed that 8/14 (57%) samples did not grow toxin-producing *C. diff*, while 6/14 (43%) samples grew toxigenic *C. diff*. All positive toxigenic culture results were concordant with RNA-NAAT and discordant with either toxin EIA or DNA-NAAT.

**Fig. 3.**
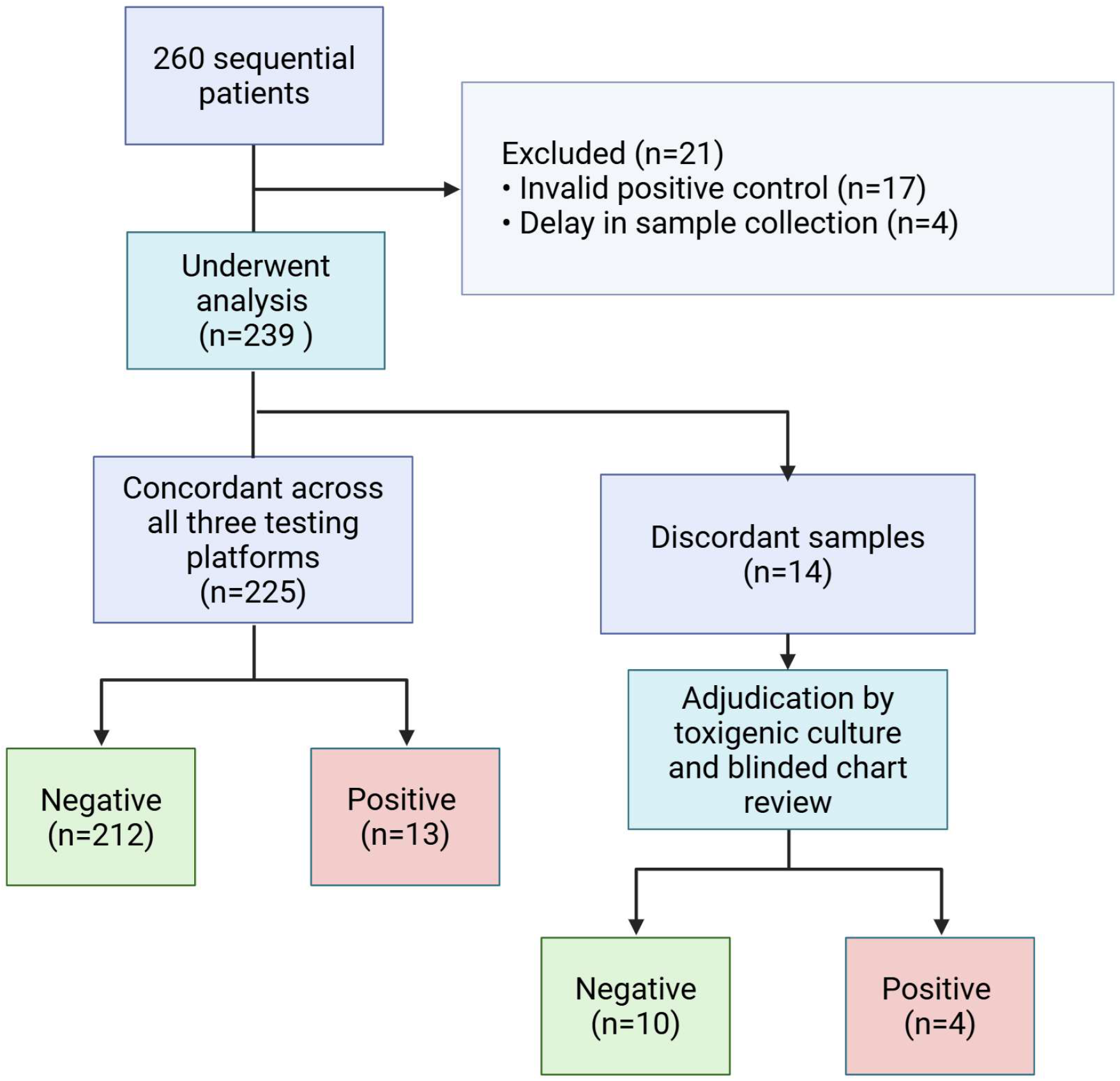
Clinical study design.

**Fig. 4.**
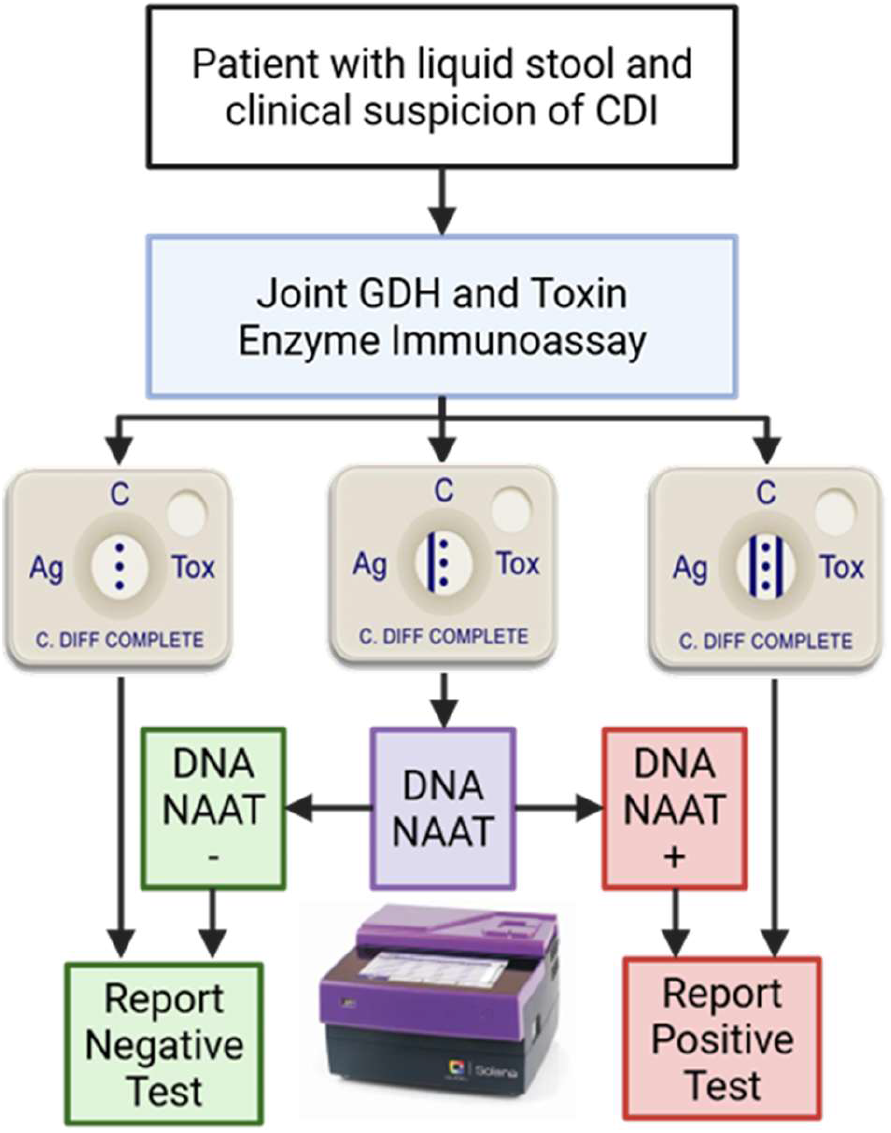
Standard of care 2-step algorithm utilized in the current study. Ag-antigen, Tox-toxin, CDI-*C. difficile* infection, GDH-Glutamate dehydrogenase, NAAT-Nucleic acid amplification test

Utilizing definitive post adjudication results (n=239), we next compared the magnitude of RNA NAAT toxin A/B ΔCt shifts in samples from patients with active toxigenic CDI (n=17) or without CDI (n=222). Patients with active CDI exhibited a statistically significant increase in toxin A/B ΔCt shifts (mean of 6.5 vs 0.07; p value 0.0002) (**Figure 5A**).

**Fig. 5.**
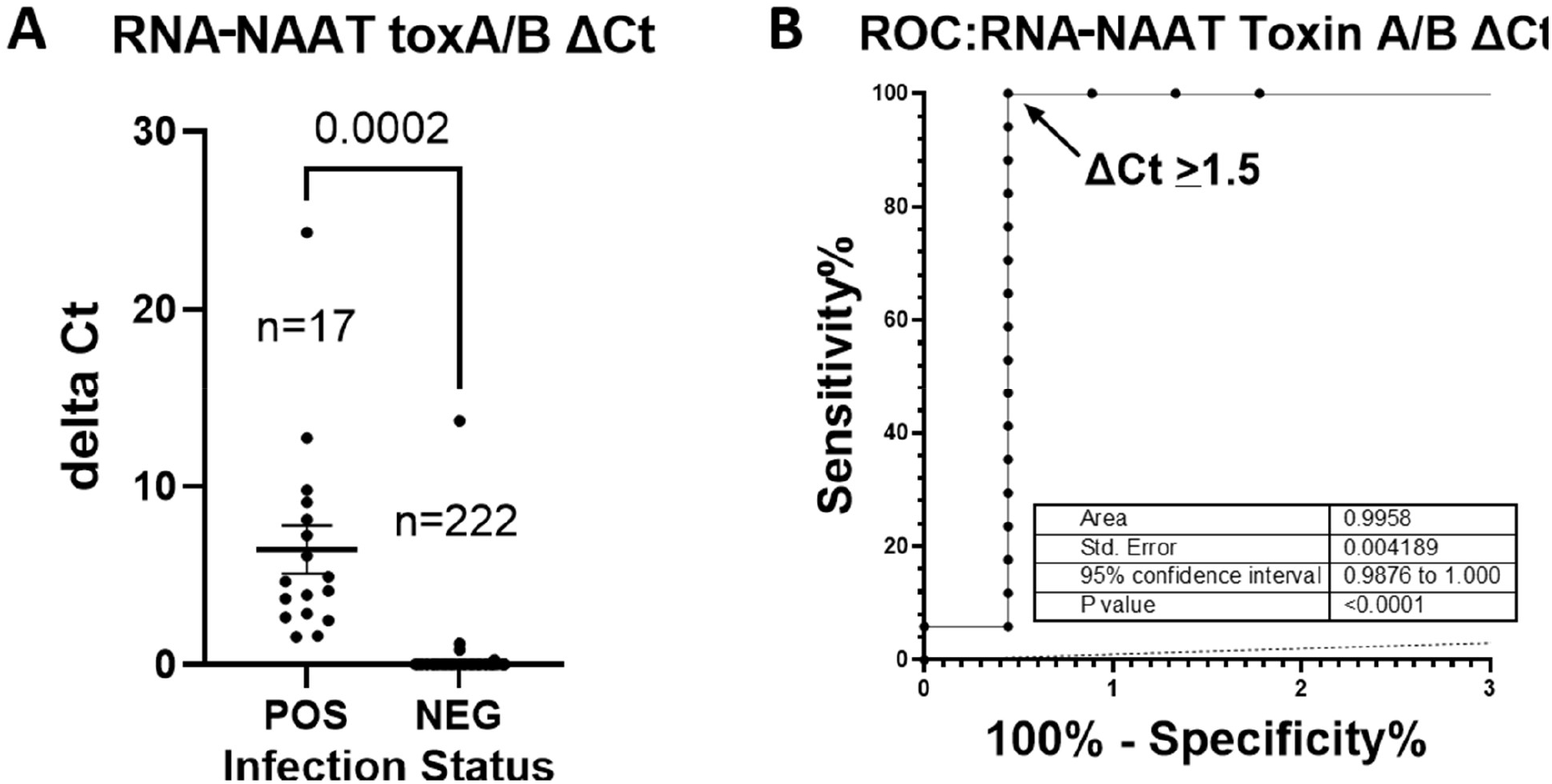
Receiver Operator Characteristic (ROC) curves identify a ΔCt ≥ 1.5 with optimal test performance characteristics. To identify an optimal RNA-NAAT delta Ct value threshold, we evaluated 239 samples with all 6 test platforms and adjudicated discordant results with toxigenic culture and blinded chart reviews. (**A)** RNA-NAAT ΔCt values for toxin A/B were plotted in patients with (n=17) or without (n=222) active CDI. An unpaired t test with Welch’s correction was used to determine statistical significance (p<0.05). (**B)** RNA-NAAT ΔCt values for toxin A/B underwent ROC curve analysis yielding an area under the curve of 0.9958 with a p value < 0.0001.

Receiver operator characteristic (ROC) curve analysis was performed to identify the RNA NAAT toxin A/B ΔCt cutoff threshold with optimal sensitivity and specificity. As shown in **Figure 5B**, the ROC curve exhibited a large area under the curve (AUC) of 0.9958 +/-0.0042, indicating excellent overall test performance. A ΔCt cutoff ≥1.5 was identified as optimal yielding a sensitivity of 100% and specificity of 99.5%. Notably, blinded chart reviews performed on three samples with RNA NAAT toxin A/B ΔCt shifts < 1.5(all toxin EIA negative and DNA-NAAT negative) were independently determined not to represent active CDI.

### RNA NAAT exhibits improved sensitivity and specificity relative to comparator assays

Diagnostic performance contingency tables for each test strategy are highlighted in **Figure 6**. RNA NAAT demonstrated excellent performance, with 100% sensitivity, 99.5% specificity, a positive predictive value (PPV) of 94.4%, and a negative predictive value (NPV) of 100% (**Figure 6A**). GDH EIA also showed 100% sensitivity but had a markedly lower specificity of 86%, resulting in a PPV of 35.4% while maintaining a NPV of 100% (**Figure 6B**). In contrast, Toxin EIA displayed 88.2% sensitivity and 98.2% specificity, with a PPV of 78.9% and an NPV of 99.1% (**Figure 6C**). DNA NAAT yielded 88.2% sensitivity, 96.8% specificity, a PPV of 68.2%, and a NPV of 99.1% (**Figure 6D**). When using the forward two-step algorithm (EIA → NAAT), sensitivity remained high at 100%, with 96.4% specificity, a PPV of 68.0%, and a NPV of 100% (**Figure 6E)**. The reverse two-step algorithm (NAAT → EIA) produced 88.2% sensitivity, 99.5% specificity, a PPV of 93.8%, and a NPV of 99.1% (**Figure 6F**). Collectively, these data highlight the trade-offs between individual test modalities and algorithmic strategies and underscore RNA NAAT’s improved specificity relative to toxin EIA and improved sensitivity relative to DNA NAATs.

**Fig. 6.**
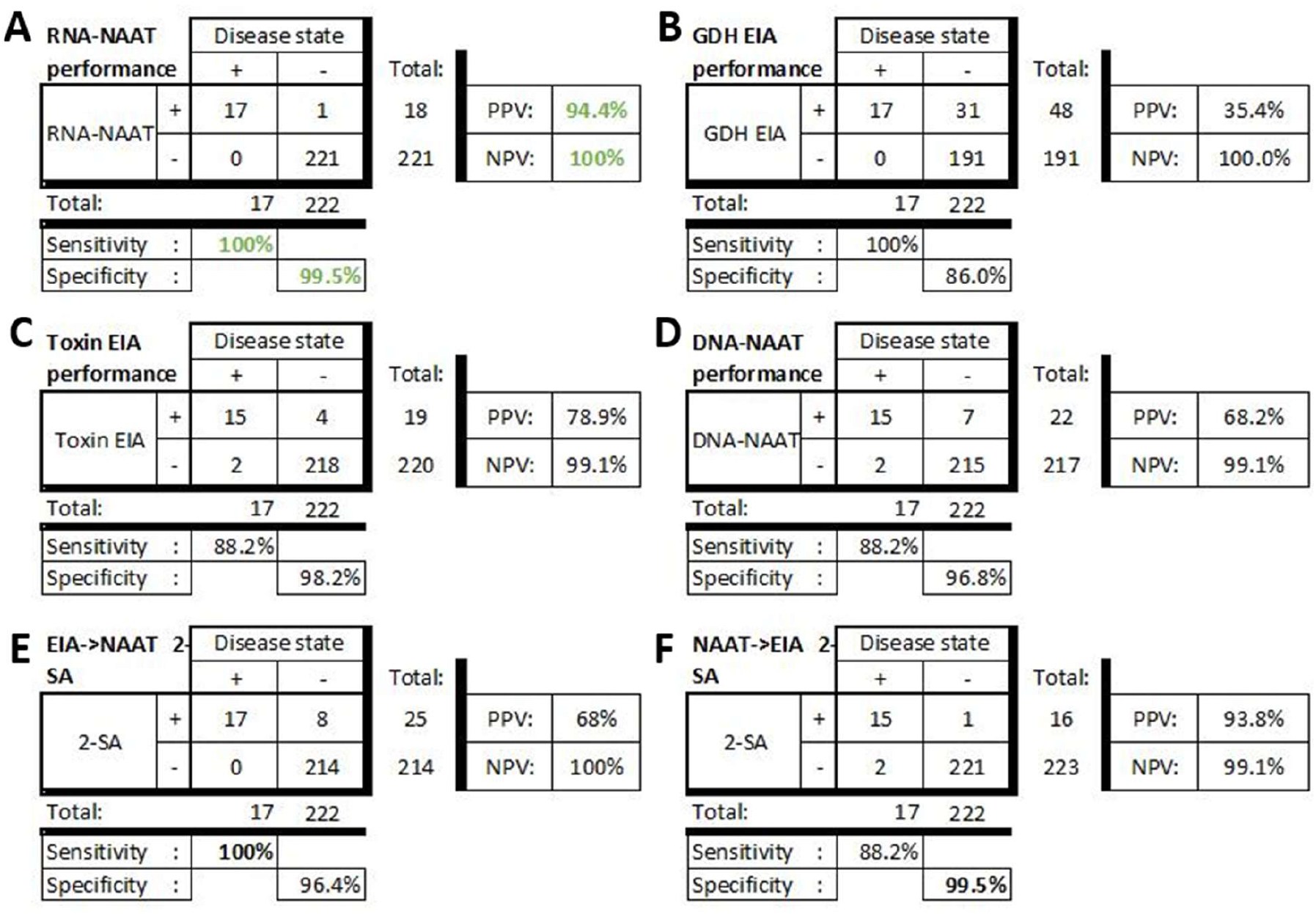
RNA NAAT exhibits increased sensitivity, specificity, positive and negative predictive values relative to comparator tests. Test results (n=239) are shown for each assay with the post-adjudication definitive disease state as the gold standard. Results are shown for (**A)** RNA-NAAT, (**B)** DNA-NAAT, (**C)** Toxin EIA, (**D)** GDH EIA, (**E)** Two Step algorithm (2-SA)-Toxin EIA→ DNA-NAAT, and **(F)** 2-SA-DNA-NAAT→ Toxin EIA.

## DISCUSSION

RNA NAAT demonstrated excellent performance, with 100% sensitivity, 99.5% specificity, a positive predictive value (PPV) of 94.4%, and a negative predictive value (NPV) of 100%. It exhibited no cross-reactivity with GI pathogens, commensal flora, or closely related species and outperformed all comparators, demonstrating the combined strength of toxin EIA (specificity 99.5% vs 98.2%) and DNA-NAAT (sensitivity 100% vs 88.2%), with markedly fewer false positives (1 vs 7).

Targeting RNA rather than DNA enhanced both clinical sensitivity and specificity for distinguishing active CDI from colonization. Typically, when the sensitivity of a test increases its specificity decreases and vice versa (*24*). However, RNA transcripts, especially rRNA, exist at far higher copy numbers than genomic DNA, enabling a lower LoD and increased analytical sensitivity (*10*).

In this study, the LoD of the 23S rRNA target was 30–70-fold lower than that of the Solana DNA NAAT, enabling more sensitive detection of *C. diff* in clinical samples (*25*). Given the flexibility to load substantially more nucleic acid extract (up to a 10-fold greater volume), future optimization may further enhance analytical sensitivity relative to DNA-NAAT, potentially approaching the ∼400-fold increase in 23S rRNA versus DNA reported by Matsuda *et al. (10*). As demonstrated in our evaluation of patient samples, this increased analytical sensitivity yielded a 100% NPV, providing practitioners with exceptional confidence to rule out CDI and focus on alternative etiologies.

Importantly, we also show that targeting toxin RNA yields increased analytical sensitivity for metabolically active, toxin-producing *C. diff* compared with toxin EIA. The Alere toxin EIA used in this study has a LoD of 0.63 ng/mL for toxin A and 0.16 ng/mL for toxin B (*27*). Although no conversion factor exists between toxin concentration and colony-forming units, we demonstrate that RNA-NAAT exhibits a LoD for toxin A/B that is 60–70-fold lower than the mean organism burden of 10^7^ CFU/mL observed in toxin EIA–positive stool samples (*28*). As noted for 23S rRNA, further optimization of reaction volumes may increase analytical sensitivity for detecting toxin synthesis. As shown in our clinical evaluation, this enhanced analytical sensitivity for toxin production resulted in a PPV of 94.4%, compared with 78.9% for toxin EIA and 68.2% for DNA NAAT, substantially improving provider confidence that a positive test reflects true infection with active *C. diff*–mediated tissue pathology.

Increased specificity relative to DNA NAAT is due primarily to the direct functional link of toxin RNA to toxin protein and tissue pathology (*29*). In this study, ΔCt shifts were observed only in metabolically active toxigenic *C. diff*. This is consistent with Senoh et al. who showed that *tcdA* RNA occurs only in vegetative cells, whereas *tcdA* DNA is detectable in spores, vegetative cells, and dead cells (*30*). Transcriptomic studies across Bacillota show major differences between vegetative cells and spores, where RNA largely reflects nucleotide storage rather than active gene expression (*31-34*). In contrast, DNA persists after cell death, allowing amplification from spores and culture-negative samples containing nonviable organisms (*30*).

GDH EIA showed 100% sensitivity but had markedly lower specificity at 86%, resulting in a PPV of 35.4% while maintaining a NPV of 100%. Comparable sensitivity of GDH and RNA NAAT in our data set is surprising as antigen tests are innately less analytically sensitive than molecular assays (*5*). Clinical chart review adjudication of discordant tests may explain this distinction in our dataset relative to other studies utilizing suboptimal “gold standard” tests (e.g. toxigenic culture). In contrast, we expected the observed decreased specificity of GDH EIA vs RNA NAAT (99.5%), given its inability to distinguish toxigenic from nontoxigenic *C. diff* and cross reactivity with closely related species (*5, 27*).

Toxin EIA exhibited 88.2% sensitivity and 98.2% specificity, with a PPV of 78.9% and a NPV of 99.1%. The decreased sensitivity of toxin EIA vs RNA NAAT (100%) was anticipated and primarily due to the increased analytical sensitivity of molecular methods vs antigen: antibody complexes (*24, 27*). Importantly, the comparable specificity for active disease (not just organism detection) of toxin EIA with RNA NAAT (99.5%) is due to the direct functional link between both toxin RNA and protein with tissue pathology.

DNA-NAAT yielded 88.2% sensitivity, 96.8% specificity, a PPV of 68.2%, and a NPV of 99.1%. The increased sensitivity of RNA NAAT relative to Solana DNA NAAT is explained by log-fold increases in RNA vs DNA targets. However, the magnitude of the drop in sensitivity relative to RNA NAAT (100%) is surprising as the literature consistently shows the sensitivity of DNA NAATs as higher than GDH and toxin EIAs, but remains within the reported 95% CI from its package insert (86.7-96.4%) (*5, 25*). Given its greater sensitivity, the inclusion of RNA NAAT resulted in two RNA NAAT positive but DNA NAAT negative samples. Both cases had appropriate risk factors (antibiotic exposure), increased stool frequency, and significant leukocytosis of >24×10^3^ cells/cm^2^, which both rapidly decreased (>50%) within 72h of starting oral vancomycin, consistent with initiation of treatment in the setting of true CDI.

In contrast, the improved specificity of RNA NAAT (99.5%) vs DNA NAAT was anticipated given the rationale described above for toxin EIA. Of the 7 false positive DNA NAATs, 3 patients had negative results reported clinically owing to the fact that the first step of the 2-SA had concordant negative GDH/toxin EIA testing. None of these patients had any evidence of CDI in the subsequent 6 months. Of the 4 tests that were reported as positive, three received substantial treatment courses, and the other patient transitioned to hospice for cervical cancer and non-*C. diff*-associated sepsis. Including the 2 doses received by the hospice patient, the average number of doses provided was 80, equivalent to twice the recommended 10-day course, with all of them having their course extended and/or empiric therapy for subsequent infection (found to be non-*C. diff*-associated) based upon the reported positive test. These four patients were all substantially immunocompromised, and all had other processes consistent with symptoms for which the test was ordered (bacteremia, candidemia, urinary tract infections, chronic medication-associated diarrhea, tube feeds). These cases illustrate the challenging landscape clinicians face when interpreting positive test results in highly complex patients and highlight the potential benefit of RNA-NAAT in providing diagnostic clarity.

The forward 2-SA (EIA → NAAT) exhibited 100% sensitivity, 96.4% specificity, a PPV of 68.0%, and a NPV of 100%. In contrast, the reverse 2-SA (NAAT → EIA) showed 88.2% sensitivity, 99.5% specificity, a PPV of 93.8%, and a NPV of 99.1%. RNA NAAT showed increased specificity (99.5%) and increased sensitivity (100%), relative to the forward and reverse algorithms, respectively. While 2-SAs are practical and offer financial benefit, the significant reduction in false positives achieved with RNA-NAAT has the potential to produce substantial system-wide savings by reducing unnecessary antibiotics, shortening hospital stays, and preventing diagnostic delays (*5, 29*).

If RNA NAAT is packaged in a relatively low cost and easy-to-use format, its superior test performance characteristics merit its implementation as a primary test for CDI. However, as a relatively complex laboratory developed test (LDT), its implementation is currently limited to higher complexity clinical labs with the infrastructure to offer and interpret custom multiplex RT-PCR assays.

At our institution, we favor the use of RNA NAAT on a specific subset of patients (7/239; 2.9%) in which toxigenic *C. diff* was detected in stool via DNA NAAT but expression of toxin A/B protein was not confirmed by EIA. This approach enables the majority of samples to be processed in the main hospital microbiology lab via the forward 2-SA, while selecting out the subset of cases that would benefit from highly sensitive detection of toxin A/B RNA via RNA NAAT, yielding fewer false positives and improved patient care.

Notably, RNA NAAT also detects binary toxin, a potentially useful prognostic indicator of severe disease, associated with worse disease and epidemic strains (*5, 20-22*). In this study, 7/17 (41%) of patients with CDI additionally tested positive for *cdtB* via RNA-NAAT. Consistent with worse disease outcomes, one binary toxin RNA^+^ patient exhibited recurrent disease spanning a 2-year interval and 3 additional patients died within four months from fulminant CDI.

A limitation of the current study is the lack of an optimal gold standard comparator test that accurately detects the presence of metabolically active toxigenic *C. diff* in patient stool samples (*5*). Given RNA NAAT’s superior test performance characteristics, it is possible that it may serve as the gold standard in future studies. In the interim, discordant results were adjudicated via a two-step blinded chart review process that is potentially susceptible to omission, misinterpretation, and subjective judgment of the patient’s clinical scenario.

As noted above, future focus areas include RNA NAAT integration into cartridge-based, sample-to-answer platforms that enable broader adoption, comprehensive analyses of cost savings relative to current diagnostic algorithms, and assessment of the prognostic value of binary toxin RNA transcripts. Although this study focuses on a single bacterial pathogen, the methods and diagnostic framework developed here have broad applicability to other pathogens, including spore-forming pathogens, for which organism detection alone is insufficient to distinguish colonization from active infection.

## MATERIALS AND METHODS

### Experimental Design

Power calculations (80% power; α = 0.05) indicated that ≥240 evaluable samples were required to detect a 5% difference in diagnostic performance. Thus, 260 sequential specimens were collected to account for potential invalid results. Samples were excluded if the RNA-preserved specimen was collected more than 24 hours after the corresponding non–RNA-preserved specimen or after initiation of oral vancomycin therapy. During assay development, RNA-NAAT was performed in triplicate. Consistent with their use in the clinical setting, RNA-NAAT and standard-of-care tests performed on patient samples were run in singlicate. All procedures using human materials or data were approved by the Institutional Review Board at the University of Alabama at Birmingham (IRB-00003200).

### *C. difficile* culture and control strains

Toxigenic (ATCC 9689) and non-toxigenic (ATCC 700057) *C. diff* strains were streaked on brain heart infusion agar supplemented with yeast extract (BHIS; BD difco) or CDC Anaerobe 5% Sheep Blood Agar Plate (BD) and incubated for 24-72 hours anaerobically at 37°C for 5 days. Single colonies were then transferred to chopped meat carbohydrate broth (CMCB; BD) overnight, before being diluted 1:100 and cultured for 6 hours to an approximate concentration of 3×10^8^ colony forming units (CFU)/mL. A total of 5mL of culture was pelleted, resuspended in 1mL RNAlater, and stored at 4°C overnight. RNA-preserved sample was centrifuged at 6000*g for 10 minutes, with the resultant pellet frozen at - 80°C until use. Toxigenic cultures were performed by culturing 100 µL of discordant sample on *C. diff*-selective cycloserine-cefoxitin fructose agar (CCFA; Thermofisher Remel) grown anaerobically at 37°C for 72 hours. Ultraviolet light was used to identify green fluorescent colonies with identities confirmed by MALDI-TOF MS (Biomerieux Vitek MS). *C. diff* colonies were then inoculated onto CMC broth and incubated for another 72 hours, prior to evaluation by Toxin EIA (Alere).

### Primer/Probe Design

All primers and probes used in the current study are outlined in **Table 1**. RNA-NAAT primers and probes targeting the 23S ribosomal RNA (rRNA) were designed to specifically detect the presence of *C. diff* with or without the ability to produce toxin. Primers and probes targeting *tcdA, tcdB*, and *cdtB*, identify RNA expression of toxin A, toxin B, and binary toxin, respectively. A positive control primer/probe set, called BactQuant, was incorporated into the RNA NAAT assay to target highly-conserved bacterial 16S rRNA sequences enabling confirmation of nucleic acid extraction from bacteria in stool. The SYBR™ Green Universal Master Mix (Applied Biosystems) was used in melt curve analyses to ensure that each primer set yielded a single amplicon. Fluorescent probes were designed to ensure compatibility with the 5 filter cubes present in the QuantStudio (QS) 5 RT-PCR system (ThermoFisher). After initial validation of individual primer specificity, probes for Toxin A and Toxin B were conjugated to the same fluorescent reporter probe (FAM) to enhance the sensitivity of detecting RNA encoding the etiologic agent of disease as well as enabling ROX reporter dye normalization to mitigate inter-assay variability. IDT PrimerQuest was used for primer design. NCBI BLAST was used for *in silico* analysis of target specificity including microbial and human genomes.

**Table 1.**
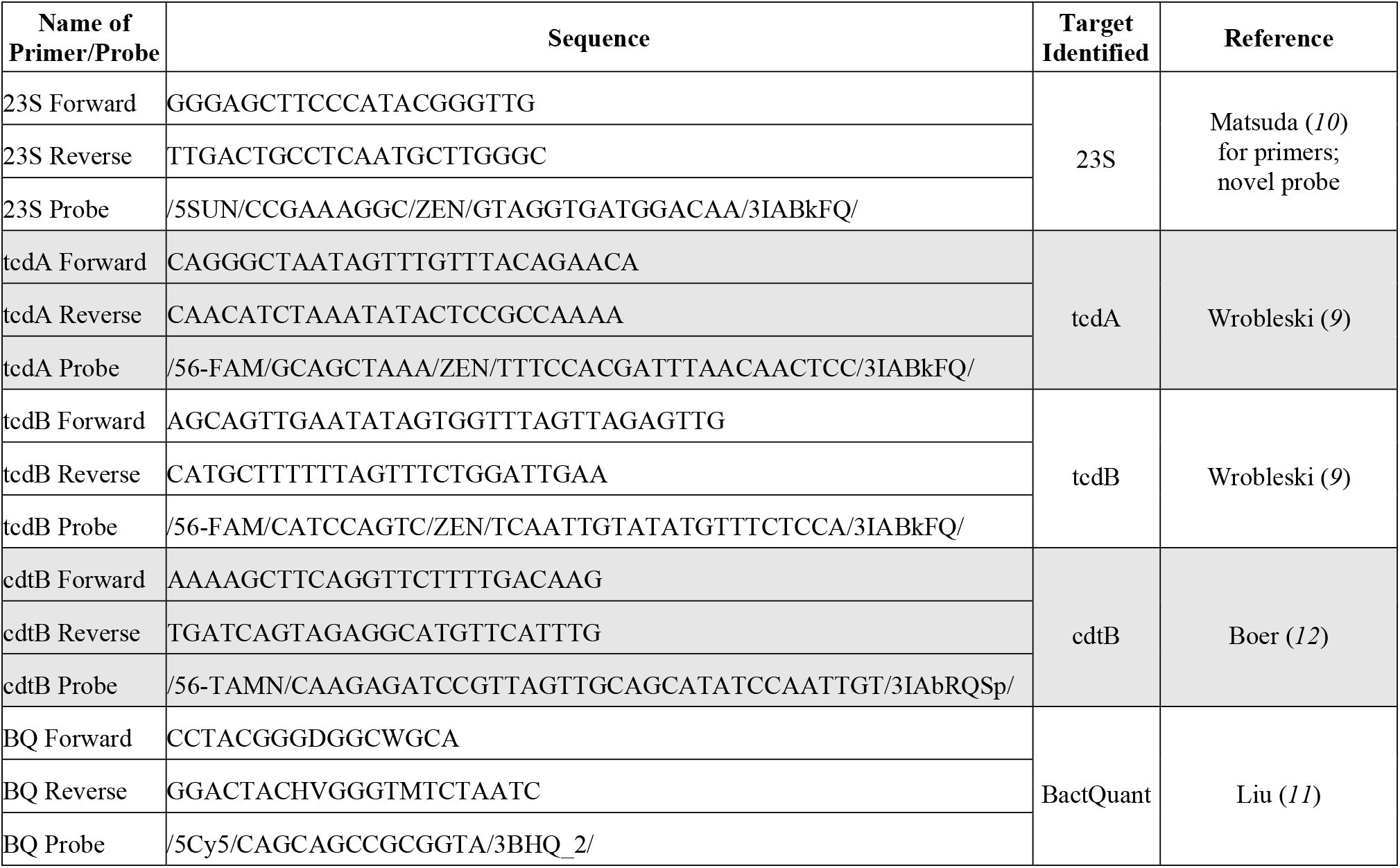
Primers and probes used to target *C. difficile*. genes of interest.

### RNA extraction and RT-PCR

Samples were homogenized in Qiagen bead bashing tubes using the Qiagen PowerLyzer 24 for 5 minutes at 2500 RPM. Homogenized samples were centrifuged at 14,000*g for 1 minute to pellet cell debris. The supernatant was processed for RNA extraction using the Promega Maxwell RSC Simply RNA Tissue Kit which includes a single DNase digestion. TaqMan PCR was carried out using TaqMan Fast Virus 1-step Master Mix (ThermoFisher). PCR primer-probe mixes were prepared fresh for each experiment with ROX used as a passive reference dye. Each extracted sample was run in duplicate; one sample was incubated at 95°C for 5 minutes to inactivate reverse transcriptase. The QS5 was used to perform RT-PCR with the following cycling parameters: Initial denaturation at 95°C for 20 seconds followed by 45 cycles of denaturation at 95°C for 3 seconds alternating with annealing/extension at 60°C for 30 seconds. For each run, thresholding was set for each target at two-fold the maximum signal of the negative control.

### RNA NAAT result interpretation

Because RNA NAAT is intended for use in patient care, we developed the test using RNA extraction and PCR platforms commonly employed for laboratory-developed tests (LDTs) in CLIA-certified clinical laboratories. Similar to other clinical extraction instruments, the Promega Maxwell Simply RNA Tissue Kit with DNase digestion, used in this study, can extract RNA from bacteria; however, complete removal of bacterial DNA from samples is not practical. Although RNA is highly enriched during this process, qualitative detection of a positive signal from toxin primer/probe sets cannot accurately distinguish between copies of the gene present in the *C. diff*. DNA genome and actively transcribed mRNA. To address this limitation and develop a practical assay suitable for implementation in a CLIA laboratory environment, we utilized heat inactivation of the reverse transcriptase enzyme. In this approach, a single RT-PCR reaction is prepared with all required amplification reagents, and the sample is divided into two wells. In one well, the RT-PCR reaction proceeds normally, allowing amplification of both the gene encoded in the DNA genome and actively transcribed mRNA captured and preserved from stool in an RNA preservative solution (eNAT swab) at the patient’s bedside. The second well undergoes the same process but includes an initial heat inactivation step, which blocks conversion of RNA to cDNA, limiting amplification to the DNA gene only. Using this paired-sample approach, an upward shift in the ΔCt value of the heat-inactivated sample indicates the presence of target RNA. Samples testing positive for the 23S rRNA gene and *toxA/B* genes were considered positive for toxigenic *C. diff*; however, only samples showing a ΔCt shift greater than 1.5 (rationale for this threshold described below) for *tcdA/tcdB* targets were deemed positive for RNA expression, indicating the presence of metabolically active toxigenic *C. diff*.

### Clinical study design

**Figure 3**. depicts the approach used to evaluate the clinical performance of RNA NAAT relative to 5 currently available commercial diagnostic strategies (3 individual tests plus two reflex algorithms). In brief, stool samples were collected from 260 sequential patients at a quaternary referral hospital (UAB Hospital, >1200 licensed beds) for whom CDI testing was ordered. Each sample was evaluated by our institutional standard of care (SOC), a forward 2-SA, consisting of a GDH/toxin EIA screen (Alere Quik Check Complete) followed by DNA-NAAT (Quidel Solana *C. diff* Assay) **(Figure 4)**.

At the same time as the SOC sample, additional samples were collected at the bedside into either a sterile container or directly into an eNAT collection device (Copan) designed to lyse cells and preserve nucleic acids. All samples were either stored at 4C and processed within 24 hours or stored at -80C for batched processing. All samples were evaluated by each test method. Upon chart review and sample analyses, a subset were excluded from further analysis due to either collection >24 hours after initiation of oral vancomycin therapy (n=4) or invalid positive controls (n=17).

### Adjudication of discordant results

All samples that were discordant across test platforms underwent toxigenic culture and clinical adjudication (n=14). Although, toxigenic culture is considered the gold standard it does not distinguish between metabolically inactive spores and vegetative bacilli. Therefore, definitive adjudication of discordant samples was determined by a two-step blinded chart review process designed to minimize bias from knowledge of reported test results. Four clinical infectious disease fellows (AJ, MP, LEH, TA) each reviewed 4-5 patient charts for discordant samples to extract relevant clinical information in a structured case format. Blinded cases were then reviewed by infectious disease fellows (3 per case) who did not have access to that patient’s chart (AJ, JK, LEH, TA). Utilizing the 2017 SHEA/IDSA Clinical Practice Guidelines for *Clostridioides difficile* Infection in Adults and Children as guidance (*35*), blinded ID fellows were then asked to determine 1) If *C. diff* testing was warranted based upon the clinical scenario, and 2) If the patient’s clinical presentation (including response to treatment) was consistent with active CDI. Each discordant result was adjudicated based on a simple majority (>/= 2/3 fellows in agreement) of responses.

## Data Availability

All data are available in the main text or the supplementary materials.

## List of Supplementary Materials

Data File S1. Ct values from all experiments with associated EIA and DNA-NAAT results

## Acknowledgments

The authors would like to thank the UAB Hospital Microbiology Laboratory, the UAB Fungal Reference Laboratory, and the UNC Hospitals Microbiology Laboratory for research technical support. Figures 1, 3, and 4 were created in BioRender, available at https://BioRender.com/j9qxvjf.

## Author contributions

Conceptualization: MBM, SML

Methodology: KDL, DJS, JH, TD, JMR

Investigation: KDL, DJS, JH, TD, DM

Clinical Adjudication: TA, LEH, AJ, MP, JK

Visualization: KDL, DJS

Funding acquisition: SML

Project administration: SML

Supervision: SML

Writing – original draft: KDL, SML

Writing – review & editing: KDL, SML, DJS, JH, TD, DM, TA, LEH, AJ, MP, JK, MBM, JMR

## Data and materials availability

All data are available in the main text or the supplementary materials.

## References and Notes

1. J. S. H. Martin, T. M. Monaghan, M. H. Wilcox, Clostridium difficile infection: epidemiology, diagnosis and understanding transmission. Nat. Rev. Gastroenterol. Hepatol. 13, 206–216 (2016).

2. F. C. Lessa et al., Burden of Clostridium difficile Infection in the United States. N. Engl. J. Med. 372, 825–834 (2015).

3. F. Prechter, K. Katzer, M. Bauer, A. Stallmach, Sleeping with the enemy: Clostridium difficile infection in the intensive care unit. Crit. Care 21, 260 (2017).

4. S. M. Brecher, S. M. Novak-Weekley, E. Nagy, Laboratory Diagnosis of Clostridium difficile Infections: There Is Light at the End of the Colon. Clin. Infect. Dis. 57, 1175–1181 (2013).

5. C.-A. D. Burnham, K. C. Carroll, Diagnosis of Clostridium difficile Infection: an Ongoing Conundrum for Clinicians and for Clinical Laboratories. Clin. Microbiol. Rev. 26, 604–630 (2013).

6. S. M. Leal et al., Quantitative Thresholds Enable Accurate Identification of Clostridium difficile Infection by the Luminex xTAG Gastrointestinal Pathogen Panel. J. Clin. Microbiol. 56, 10.1128/jcm.01885-01817 (2018).

7. S. R. Curry et al., Natural History of Clostridioides difficile Colonization and Infection Following New Acquisition of Carriage in Healthcare Settings: A Prospective Cohort Study. Clin. Infect. Dis. 77, 77–83 (2023).

8. M. Gilboa, N. Baharav, E. Melzer, G. Regev-Yochay, D. Yahav, Screening for Asymptomatic Clostridioides difficile Carriage Among Hospitalized Patients: A Narrative Review. Infect. Dis. Ther. 12, 2223–2240 (2023).

9. D. Wroblewski et al., Rapid Molecular Characterization of Clostridium difficile and Assessment of Populations of C. difficile in Stool Specimens. J. Clin. Microbiol. 47, 2142–2148 (2009).

10. K. Matsuda et al., Sensitive Quantification of Clostridium difficile Cells by Reverse Transcription-Quantitative PCR Targeting rRNA Molecules. Appl. Environ. Microbiol. 78, 5111–5118 (2012).

11. C. M. Liu et al., BactQuant: An enhanced broad-coverage bacterial quantitative real-time PCR assay. BMC Microbiol. 12, 56 (2012).

12. R. F. d. Boer et al., Evaluation of a rapid molecular screening approach for the detection of toxigenic Clostridium difficile in general and subsequent identification of the tcdC Δ117 mutation in human stools. J. Microbiol. Methods 83, 59–65 (2010).

13. C. R. Polage et al., Overdiagnosis of Clostridium difficile Infection in the Molecular Test Era. JAMA Intern. Med. 175, 1792–1801 (2015).

14. G. R. Madden, D. C. Smith, M. D. Poulter, C. D. Sifri, Propensity-Matched Cost of Clostridioides difficile Infection Overdiagnosis. Open Forum Infect. Dis. 8, ofaa630 (2020).

15. M. Kachrimanidou et al., A two-step approach improves the diagnosis of Clostridium difficile infection. J. Microbiol. Methods 143, 17–19 (2017).

16. C. C. Bettger, S. E. Giancola, R. J. Cybulski, J. F. Okulicz, A. E. Barsoumian, Evaluation of a two step testing algorithm to improve diagnostic accuracy and stewardship of Clostridioides difficile infections. BMC Res. Notes 16, 172 (2023).

17. L. Dbeibo et al., Two-step algorithm-based Clostridioides difficile testing as a tool for antibiotic stewardship. Clin. Microbiol. Infect. 29, 798.e791-798.e794 (2023).

18. A. Guh et al., 1665. Potential underreporting of treated patients using a Clostridioides difficile testing algorithm that screens with a nucleic acid amplification test. Open Forum Infect. Dis. 9, ofac492.131 (2022).

19. E. E. Hilt, B. P. Vaughn, A. L. Galdys, M. D. Evans, P. Ferrieri, Impact of the Reverse 2-Step Algorithm for Clostridioides difficile Testing in the Microbiology Laboratory on Hospitalized Patients. Open Forum Infect. Dis. 11, ofae244 (2024).

20. D. N. Gerding, S. Johnson, M. Rupnik, K. Aktories, Clostridium difficile binary toxin CDT. Gut Microbes 5, 15–27 (2014).

21. C. Eckert et al., Prevalence and pathogenicity of binary toxin–positive Clostridium difficile strains that do not produce toxins A and B. N. Microbes N. Infect. 3, 12–17 (2015).

22. M. K. Young et al., Binary Toxin Expression by Clostridioides difficile Is Associated With Worse Disease. Open Forum Infect. Dis. 9, ofac001 (2022).

23. Microbiologics, in Microbiologics. . (2025).

24. N. Rifai, A. R. Horvath, C. Wittwer, Tietz textbook of clinical chemistry and molecular diagnostics. (Elsevier, St. Louis, Missouri, ed. Sixth edition., 2018), pp. xx, 1867 pages.

25. Q. Corporation, in Quidel Ortho Technical Documents. (2025).

26. A. Forootan et al., Methods to determine limit of detection and limit of quantification in quantitative real-time PCR (qPCR). Biomol. Detect. Quantif. 12, 1–6 (2017).

27. I. TECHLAB. (2021), chap. 16.

28. L.-L. Dionne et al., Correlation between Clostridium difficile Bacterial Load, Commercial Real-Time PCR Cycle Thresholds, and Results of Diagnostic Tests Based on Enzyme Immunoassay and Cell Culture Cytotoxicity Assay. J. Clin. Microbiol. 51, 3624–3630 (2013).

29. S. D. Bella et al., Clostridioides difficile infection: history, epidemiology, risk factors, prevention, clinical manifestations, treatment, and future options. Clin. Microbiol. Rev. 37, e00135–00123 (2024).

30. M. Senoh et al., Reverse transcription polymerase chain reaction‐based method for selectively detecting vegetative cells of toxigenic Clostridium difficile. Microbiol. Immunol. 58, 615–620 (2014).

31. B. Byrd et al., Levels and Characteristics of mRNAs in Spores of Firmicute Species. J. Bacteriol. 203, e00017–00021 (2021).

32. D. L. Craft et al., Analysis of 5′-NAD capping of mRNAs in dormant spores of Bacillus subtilis. FEMS Microbiol. Lett. 367, fnaa143 (2020).

33. G. Korza et al., Analysis of the mRNAs in Spores of Bacillus subtilis. J. Bacteriol. 201, 10.1128/jb.00007-00019 (2019).

34. G. Korza, B. Setlow, L. Rao, Q. Li, P. Setlow, Changes in Bacillus Spore Small Molecules, rRNA, Germination, and Outgrowth after Extended Sublethal Exposure to Various Temperatures: Evidence that Protein Synthesis Is Not Essential for Spore Germination. J. Bacteriol. 198, 3254–3264 (2016).

35. L. C. McDonald et al., Clinical Practice Guidelines for Clostridium difficile Infection in Adults and Children: 2017 Update by the Infectious Diseases Society of America (IDSA) and Society for Healthcare Epidemiology of America (SHEA). Clin. Infect. Dis. 66, e1–e48 (2018).

